# Monoclonal antibodies therapy for Covid-19 - Experiences at a regional hospital

**DOI:** 10.1101/2021.09.22.21263488

**Authors:** Judith Pannier, Norbert Nass, Gerhard Behre

## Abstract

Applying monoclonal antibodies against Covid-19 is a promising treatment option for avoiding severe outcomes. However, real life data, especially in regional hospitals are still scarce. We here report on our first results with this therapy in a retrospective, observational study. Indeed, compared to a risk-factor matched reference group, hospitalisation time was reduced but survival rate and kinetics of SARS-CoV-2 RT-PCR results remained apparently unaffected.

## Introduction

With the occurrence of Covid-19, a demand for the rapid development of modern approachesappeared and techniques used in oncology such as mRNA vaccination and humanized monoclonal antibodies (Buss et al. 2012) have been rapidly adapted for the use in this infectious disease (Alvi et al. 2020). The effectiveness of the monoclonal antibody (mab) cocktails Bamlanivimab/Etesevimab (Dougan et al. 2021) and Casirivimab/Imdevimab (Weinreich et al. 2021) has recently been demonstrated for patients at risk for unfavourable outcome (Taylor et al. 2021) of Covid-19 before severe symptoms appear. These mabs are directed against the SARS-CoV2-Spike Protein (Hurt et al. 2021), which facilitates the binding to the cell surface receptor ACE2. Such antibodies are either directed against highly conserved epitopes or applied in pairs to reduce the selection of S-protein mutations that would cause the development of resistant viral strains (Esmaeilzadeh et al. 2021). We here report on initial experiences with this therapy in our regional hospital.

## Methods

### Patients and ethical statement

Resident physicians and patients were informed about the monoclonal antibody (mab) therapy by mail and the media. A Covid-19 ambulance was established to survey and select patients eligible for the therapy. Patients were included within 7 days after symptom onset and positive SARS-CoV-2-RT-PCR based diagnosis as well as the presence of multiple risk factors. 34 patients were treated by either bamlanivimab alone (5) or casirivimab plus imdevimab in combination (29) from March to June 2021. Data from a reference group matched for age, sex and risk factors, and hospitalised for Covid-19 before the mab therapy became available from January to May 2021 were collected for comparison. Both patient groups were treated, applying the same guidelines. Data on symptoms, risk factors and SARS-CoV2-RT-PCR as well as routine clinical data were collected and statistically analysed as indicated. Patients were informed about this study and gave consent for the use of their data for research purposes.

### Mab treatment

Monoclonal antibodies were provided by an initiative of the German Federal Ministry of Health. The antibodies were administered as intravenous infusion over 1 hour followed by an observational period of another hour.

### Statistics

Significance of dichotomized data was determined by cross tabulations applying Fisher’s exact test or ordinal by ordinal correlation. Student’s T-test was applied for comparison of means of metric data. All calculations were dome using SPSS (IBM Corp. Released 2015. IBM SPSS Statistics for Windows, Version 23.0. Armonk, NY: IBM Corp.)

## Results

The two groups did not differ significantly in most of the clinical data such as age, sex and Covid-19 risk factors (Tab. 1 and supplementary table 1). However, there were differences in the observed symptoms, especially the occurrence of coughing and dyspnoea and, consequently, arterial pO_2_ and pCO_2_ values, which correlates with the dissimilar time between initial positive RT-PCR result, symptom onset and time of hospitalization. The most important significant difference in the two groups was found for the time of hospitalization (9.3 days versus 17.4 days, p < 0.001) but not death after positive PCR based Covid-19 diagnosis (two target amplification). Also, we found no difference in the course of the Ct-values of SARS-CoV-2 RT-PCR during hospitalisation (Fig. 1)

**Table 1:**
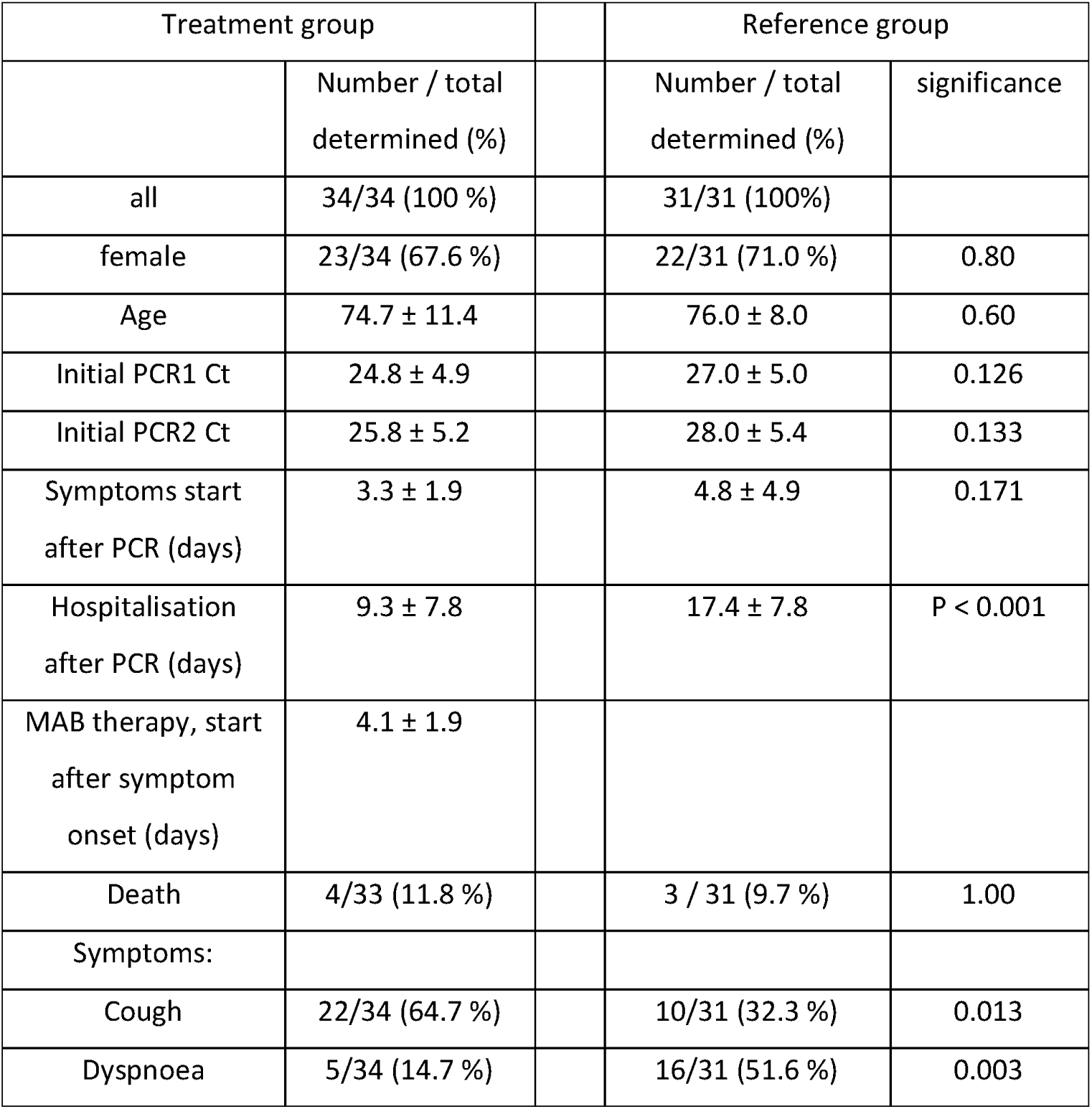
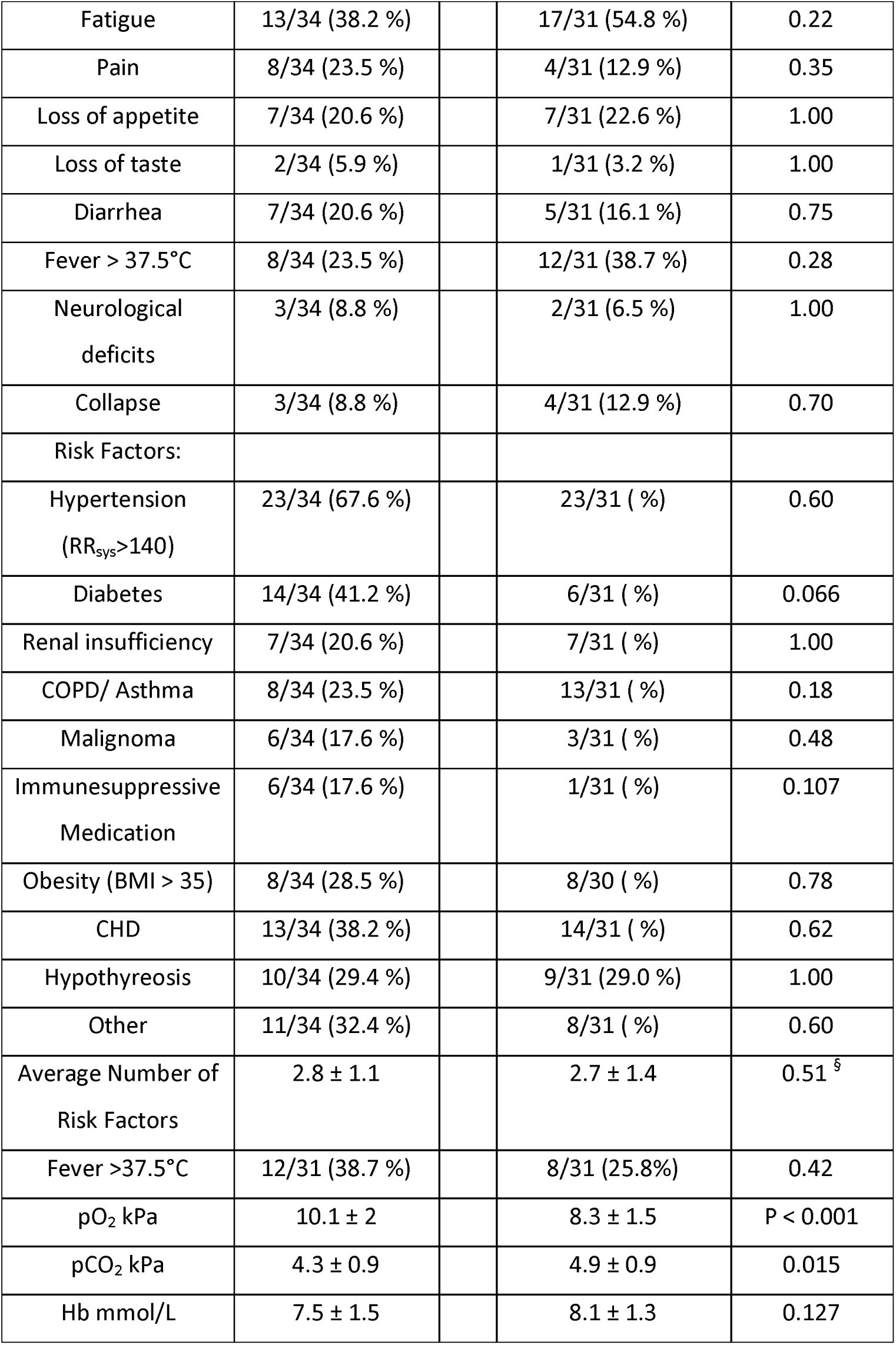
Characteristics of the study cohort. Significance for cross tabulation tests was determined by Fisher’s exact test or ordinal by ordinal correlation (§). Student’s T-test was applied for comparison of means of metric data. Mean and standard deviation are given.

| Treatment group |  | Reference group |  |
| --- | --- | --- | --- |
|  | Number / total determined (%) | Number / total determined (%) | significance |
| all | 34/34 (100 %) | 31/31 (100%) |  |
| female | 23/34 (67.6 %) | 22/31 (71.0 %) | 0.80 |
| Age | 74.7 ± 11.4 | 76.0 ± 8.0 | 0.60 |
| Initial PCR1 Ct | 24.8 ± 4.9 | 27.0 ± 5.0 | 0.126 |
| Initial PCR2 Ct | 25.8 ± 5.2 | 28.0 ± 5.4 | 0.133 |
| Symptoms start after PCR (days) | 3.3 ± 1.9 | 4.8 ± 4.9 | 0.171 |
| Hospitalisation after PCR (days) | 9.3 ± 7.8 | 17.4 ± 7.8 | P < 0.001 |
| MAB therapy, start after symptom onset (days) | 4.1 ± 1.9 |  |  |
| Death | 4/33 (11.8 %) | 3 / 31 (9.7 %) | 1.00 |
| Symptoms: |  |  |  |
| Cough | 22/34 (64.7 %) | 10/31 (32.3 %) | 0.013 |
| Dyspnoea | 5/34 (14.7 %) | 16/31 (51.6 %) | 0.003 |

|  |  |  |  |  |
| --- | --- | --- | --- | --- |
| Fatigue | 13/34 (38.2 %) |  | 17/31 (54.8 %) | 0.22 |
| Pain | 8/34 (23.5 %) |  | 4/31 (12.9 %) | 0.35 |
| Loss of appetite | 7/34 (20.6 %) |  | 7/31 (22.6 %) | 1.00 |
| Loss of taste | 2/34 (5.9 %) |  | 1/31 (3.2 %) | 1.00 |
| Diarrhea | 7/34 (20.6 %) |  | 5/31 (16.1 %) | 0.75 |
| Fever > 37.5°C | 8/34 (23.5 %) |  | 12/31 (38.7 %) | 0.28 |
| Neurological<br>deficits | 3/34 (8.8 %) |  | 2/31 (6.5 %) | 1.00 |
| Collapse | 3/34 (8.8 %) |  | 4/31 (12.9 %) | 0.70 |
| Risk Factors: |  |  |  |  |
| Hypertension<br>(RR<sub>sys</sub>>140) | 23/34 (67.6 %) |  | 23/31 ( %) | 0.60 |
| Diabetes | 14/34 (41.2 %) |  | 6/31 ( %) | 0.066 |
| Renal insufficiency | 7/34 (20.6 %) |  | 7/31 ( %) | 1.00 |
| COPD/ Asthma | 8/34 (23.5 %) |  | 13/31 ( %) | 0.18 |
| Malignoma | 6/34 (17.6 %) |  | 3/31 ( %) | 0.48 |
| Immunosuppressive<br>Medication | 6/34 (17.6 %) |  | 1/31 ( %) | 0.107 |
| Obesity (BMI > 35) | 8/34 (28.5 %) |  | 8/30 ( %) | 0.78 |
| CHD | 13/34 (38.2 %) |  | 14/31 ( %) | 0.62 |
| Hypothyreosis | 10/34 (29.4 %) |  | 9/31 (29.0 %) | 1.00 |
| Other | 11/34 (32.4 %) |  | 8/31 ( %) | 0.60 |
| Average Number of<br>Risk Factors | 2.8 ± 1.1 |  | 2.7 ± 1.4 | 0.51<sup>§</sup> |
| Fever >37.5°C | 12/31 (38.7 %) |  | 8/31 (25.8%) | 0.42 |
| pO<sub>2</sub> kPa | 10.1 ± 2 |  | 8.3 ± 1.5 | P < 0.001 |
| pCO<sub>2</sub> kPa | 4.3 ± 0.9 |  | 4.9 ± 0.9 | 0.015 |
| Hb mmol/L | 7.5 ± 1.5 |  | 8.1 ± 1.3 | 0.127 |

**Figure 1:**
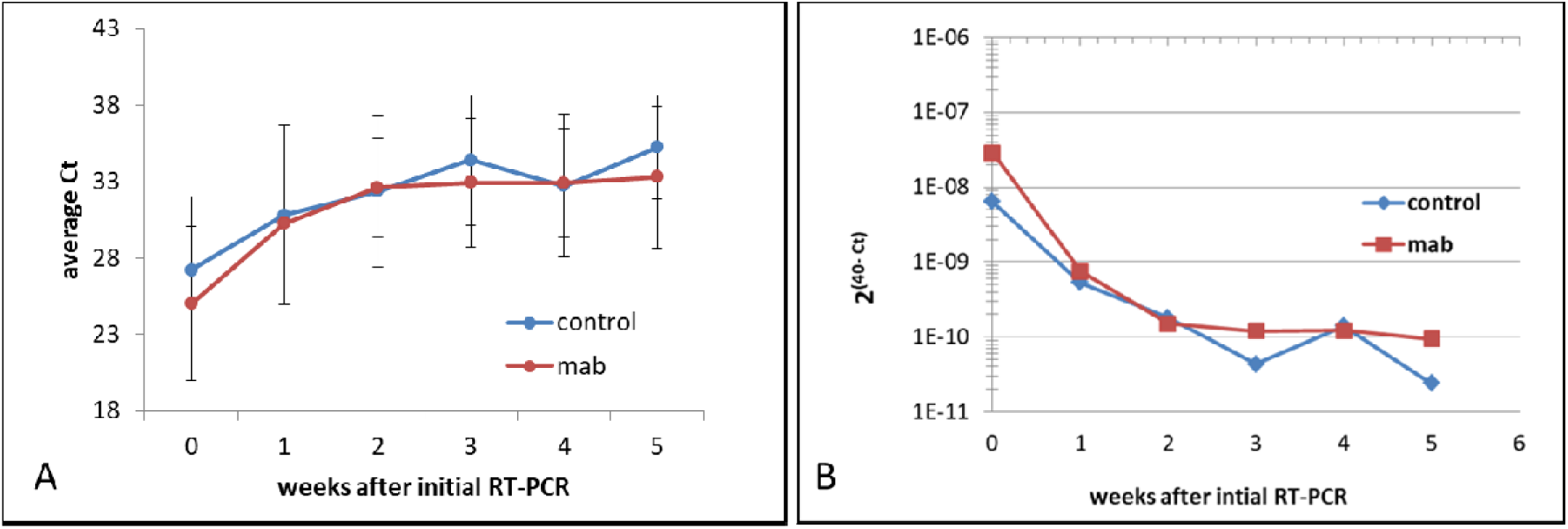
SARS-CoV-2 RT-PCR results from nasal swaps of the patients. Data are presented as average C_t_ including standard deviation (A) or 2^(40-Ct)^ (B) after initial positive RT-PCR diagnosis to visualize the decrease of viral load over time. Ct-values considered as negative were set to 40.

## Discussion

In this small retrospective observational study, we could detect a significant shortening of hospitalisation time in the mab-treated cohort, consistent with larger studies published recently (Dougan et al. 2021, Weinreich et al. 2021). This demonstrates that this treatment can be highly effective as long as patients at risk are identified early, before severe symptoms occur. This will become important to avoid a high burden of hospitalized patients when greater Covid-19 incidences will appear. To achieve this, we have encouraged residential physicians by mail and media to immediately direct their SARS-CoV-2 positive patients having significant risk factors and beginning symptoms to our hospital for early therapy. The observed differences in blood gases as well as the frequency of coughing and dyspnoe reflect the different time after first positive RT-PCR and hospitalisation. However, we were not able to reproduce the reported early decrease in RT-PCR Ct-values (Dougan et al. 2021) in the treatment group (Fig. 1). However, especially at longer times after initial PCR, only patients being still positive by RT-PCR remain in this analysis, which might skew the result. Clearly, a higher number of patients will be needed to clarify this result.

It was also disappointing that about 10 % of the patients had a fatal outcome in both groups. This result might be due to the selection of patients, having a high number of risk factors. Again, larger numbers are needed to clarify this result.

Altogether, our experiences with this mab therapy are very encouraging in terms of avoiding long hospitalisation times in risk-patients as soon as the mabs are applied early in disease progression, which is consistent with the results of larger studies published recently. Nevertheless, efficient ways to inform patients on the availability of this therapy are needed.

## Supporting information

Supplement Strobe Checklist

## Data Availability

All necessary data are included in the manuscript. Further details will be made available on reasonable request

## Acknowledgments

We thank all patients for their participation in this study.

## Author contributions

J.P. and G.B: examination and treatment of patients, data collection, manuscript editing, project administration, N.N: data collection and statistics, manuscript writing and editing.

---

**Suppl. table 1:**
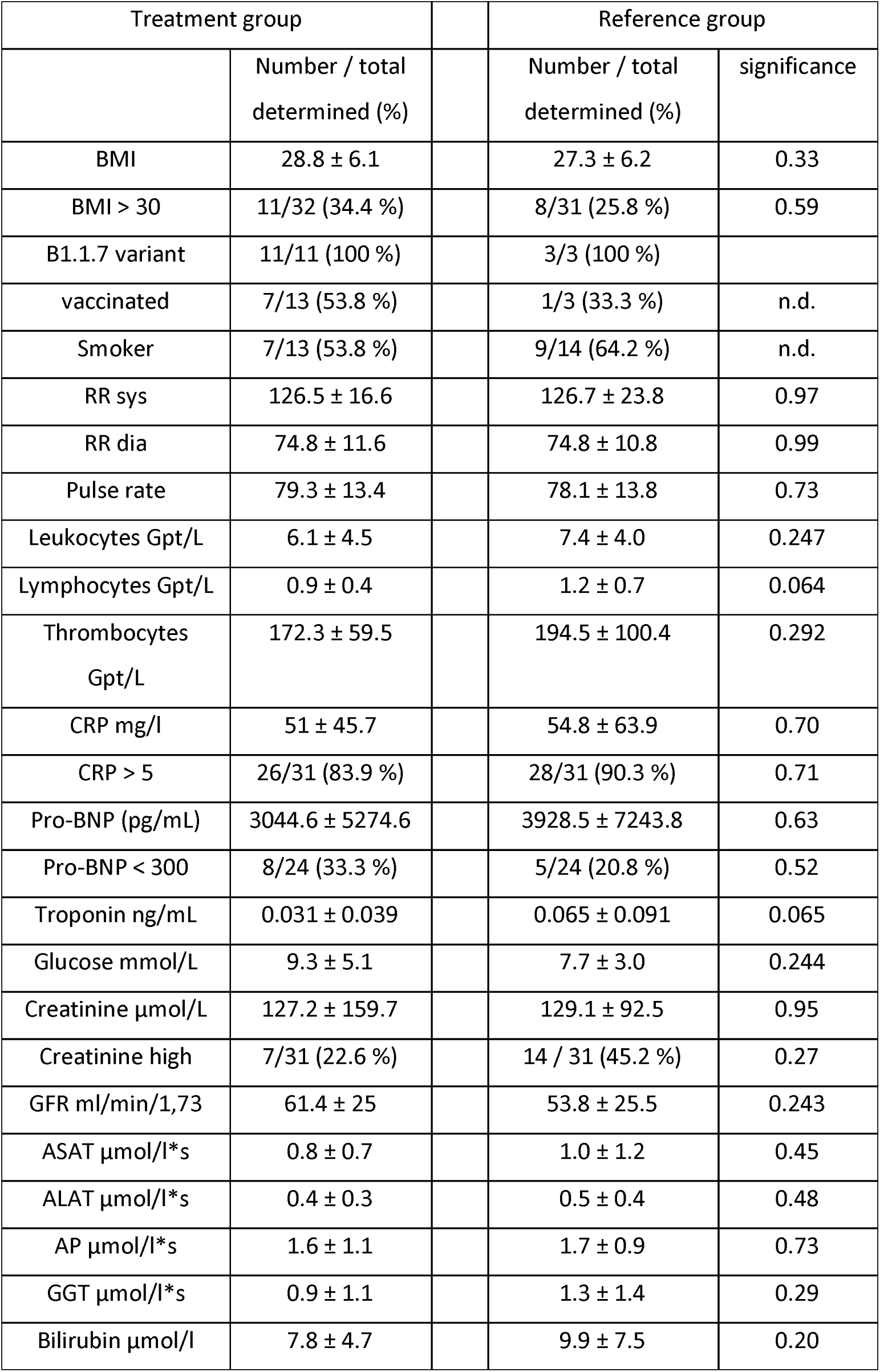
Further characteristics of the study cohort. Significance for cross tabulations was determined by Fisher’s exact test or ordinal by ordinal correlation (§). Student’s T-test was applied for comparison of means of metric data. Mean and standard deviation are given. N.d.: not determined. Supplement Strobe Checklist

| Treatment group |  |  | Reference group |  |
| --- | --- | --- | --- | --- |
|  | Number / total<br>determined (%) |  | Number / total<br>determined (%) | significance |
| BMI | 28.8 ± 6.1 |  | 27.3 ± 6.2 | 0.33 |
| BMI > 30 | 11/32 (34.4 %) |  | 8/31 (25.8 %) | 0.59 |
| B1.1.7 variant | 11/11 (100 %) |  | 3/3 (100 %) |  |
| vaccinated | 7/13 (53.8 %) |  | 1/3 (33.3 %) | n.d. |
| Smoker | 7/13 (53.8 %) |  | 9/14 (64.2 %) | n.d. |
| RR sys | 126.5 ± 16.6 |  | 126.7 ± 23.8 | 0.97 |
| RR dia | 74.8 ± 11.6 |  | 74.8 ± 10.8 | 0.99 |
| Pulse rate | 79.3 ± 13.4 |  | 78.1 ± 13.8 | 0.73 |
| Leukocytes Gpt/L | 6.1 ± 4.5 |  | 7.4 ± 4.0 | 0.247 |
| Lymphocytes Gpt/L | 0.9 ± 0.4 |  | 1.2 ± 0.7 | 0.064 |
| Thrombocytes<br>Gpt/L | 172.3 ± 59.5 |  | 194.5 ± 100.4 | 0.292 |
| CRP mg/l | 51 ± 45.7 |  | 54.8 ± 63.9 | 0.70 |
| CRP > 5 | 26/31 (83.9 %) |  | 28/31 (90.3 %) | 0.71 |
| Pro-BNP (pg/mL) | 3044.6 ± 5274.6 |  | 3928.5 ± 7243.8 | 0.63 |
| Pro-BNP < 300 | 8/24 (33.3 %) |  | 5/24 (20.8 %) | 0.52 |
| Troponin ng/mL | 0.031 ± 0.039 |  | 0.065 ± 0.091 | 0.065 |
| Glucose mmol/L | 9.3 ± 5.1 |  | 7.7 ± 3.0 | 0.244 |
| Creatinine µmol/L | 127.2 ± 159.7 |  | 129.1 ± 92.5 | 0.95 |
| Creatinine high | 7/31 (22.6 %) |  | 14 / 31 (45.2 %) | 0.27 |
| GFR ml/min/1,73 | 61.4 ± 25 |  | 53.8 ± 25.5 | 0.243 |
| ASAT µmol/l*s | 0.8 ± 0.7 |  | 1.0 ± 1.2 | 0.45 |
| ALAT µmol/l*s | 0.4 ± 0.3 |  | 0.5 ± 0.4 | 0.48 |
| AP µmol/l*s | 1.6 ± 1.1 |  | 1.7 ± 0.9 | 0.73 |
| GGT µmol/l*s | 0.9 ± 1.1 |  | 1.3 ± 1.4 | 0.29 |
| Bilirubin µmol/l | 7.8 ± 4.7 |  | 9.9 ± 7.5 | 0.20 |

## References

Alvi MM, Sivasankaran S and Singh M. Pharmacological and non-pharmacological efforts at prevention, mitigation, and treatment for COVID-19. Drug Target. 2020; 28:742–754. doi: 10.1080/1061186X.2020.1793990.

Buss NAPS, Henderson SJ, McFarlane M, Shenton JM, de Haan L. Monoclonal antibody therapeutics: history and future. Curr Opin Pharmacol. 2012; 12: 615–622 doi: 10.1016/j.coph.2012.08.001.

Dougan M, Nirula A, Azizad M, Mocherla B, Gottlieb RL, Chen P et al.et al. Bamlanivimab plus Etesevimab in Mild or Moderate Covid-19. N Engl J Med. 2021; NEJMoa2102685. doi: 10.1056/NEJMoa2102685.

Esmaeilzadeh A, Rostami S, Yeganeh PM, Tahmasebi S, Ahmadi M. Recent advances in antibody based immunotherapy strategies for COVID-19. J Cell Biochem. 2021; doi: 10.1002/jcb.30017

Hurt AC, Wheatley AK. Neutralizing Antibody Therapeutics for COVID-19. Viruses. 2021; 13: 628. doi: 10.3390/v13040628

Taylor EH, Marson EJ, Elhadi M, Macleod KDM, Yu YC, Davids R et al. Factors associated with mortality in patients with COVID-19 admitted to intensive care: a systematic review and meta131 analysis. Anaesthesia. 2021; 76: 1224–1232. doi: 10.1111/anae.15532.

Weinreich DM, Sivapalasingam S, Norton T, Ali S, Gao H, Bhore R et al. REGN-COV2, a Neutralizing Antibody Cocktail, in Outpatients with Covid-19. N Engl J Med. 2021; 384: 238–251. doi: 10.1056/NEJMoa2035002

